# Pulse pressure: effective predictor for clinical outcomes after thrombectomy

**DOI:** 10.1101/2024.11.04.24316726

**Authors:** Jianru Li, Shandong Jiang, Peizheng Guo, Yuan yuan, Jun Yu, Liang Xu, Xu Li, Xianyi Chen, Bing Fang, Zhongju Tan, Jing Xu, Cong Qian

## Abstract

**Background:** Previous guidelines for post-operative blood pressure (BP) management have focused on SBP and DBP in stroke patients who have undergone intravenous thrombolysis (IVT). Whether pulse pressure (PP) affects the prognosis of patients with ischemic stroke after endovascular thrombectomy (EVT) remains an unresolved issue.

**Methods:** In this study, we systematically collected twelve BP parameters within 24 hours after thrombectomy and regularly followed up to assess the mRS scores. We utilized univariate and multivariate logistic regression analyses to identify predictive factors for poor prognosis and morality following EVT. Restrictive cubic splines (RCS) are used to evaluate the dose-effect relationship between PP and outcome events. Subgroup analyses were conducted to assess the prognostic efficacy of mean PP across different patient groups, with a favorable outcome defined as a modified Rankin Scale (mRS) score between 0 and 3 at three months post-EVT.

**Results:** Post-EVT SBP data were available for 587/826 patients. Mean PP demonstrates a significant positive dose-response relationship with the occurrence of functional outcomes, sICH, and mortality after EVT. The predictive power and strength of association of mean PP with prognosis are superior to those of single SBP or DBP alone with the strongest ORs and highest diagnostic performance. Mean PP exhibits a linear relationship with all other outcome events, except for mortality at 12 months post-EVT.

**Conclusion:** The mean PP within 24 hours after EVT is an independent risk factor for sICH, prognosis, and mortality in stroke patients, with a stronger association and diagnostic performance than either SBP or DBP. Achieving consistent long-term control of SBP and PP may be beneficial for improving the prognosis of ischemic stroke patients.

*What is already known on this topic:* It has been confirmed that the post-procedure blood pressure (BP) are closely related to the onset and progression of stroke. Previous guidelines for postoperative blood pressure (BP) management have focused on SBP and DBP in stroke patients who have undergone intravenous thrombolysis (IVT). Whether pulse pressure (PP) affects the prognosis of patients with ischemic stroke after endovascular thrombectomy (EVT) remains an unresolved issue.

*What this study adds:* Mean PP demonstrates a significant positive dose-response relationship with the occurrence of functional outcomes, sICH, and mortality after thrombectomy. The predictive power and strength of association of mean PP with prognosis are superior to those of single SBP or DBP alone with the strongest ORs and highest diagnostic performance (AUC=0.661, 95% CI 0.617 to 0.705).

*How this study might affect research, practice or policy:* In Clinical, we usually focus on controlling post-procedure SBP or DBP after EVT. In this study, we demonstrated that the mean PP within 24 hours after EVT is an independent risk factor for sICH, prognosis, and mortality in stroke patients. Achieving consistent long-term control of SBP and PP may be beneficial for improving the prognosis of ischemic stroke patients.

## Introduction

Large vessel occlusions (LVO) account for approximately one third of acute ischemic stroke (AIS) but contribute to more than half of all stroke-related disability and mortality^1^. Since 2016, endovascular thrombectomy (EVT), have been an established treatment for patients with AIS caused by LVO^2^. Despite achieving successful recanalization (eTICI=2B or 3) after the end of the endovascular procedure, approximately half of the patients die or are left sever permanently disabled (mRS 4-6)^3,4^. Recent evidences suggested that hemodynamic management may play an important role in post-EVT care^5^. Up to 80% patients with AIS are usually accompanied with elevated blood pressure (BP) reaction after procedure, it takes a few days to return to baseline levels^6^. When ischemia occurs, the blood flow regulatory capacity of the brain tissue in the infarct area is diminished, thereby amplifying the impact of peripheral systemic BP on intracranial pressure and perfusion^7^. Raised BP and its variability post-EVT have been implicated in poor functional outcomes^8^, cerebral edema, and symptomatic hemorrhagic transformation^9^. Despite recently completed several clinical trials on BP managements post-EVT, the conclusions remain inconsistent, and there are still controversies over whether implement lowering BP (<140 mmHg) is effective in improving prognosis^10^, or reducing the occurrence of spontaneous cerebral hemorrhage^11^, or when to implement intensive BP lowering therapy^12^.

These above studies primarily focused on the association between postoperative SBP and outcome events. LEE et al. found that PP seemed to be a better prognostic indicator than SBP and mean arterial pressure (MAP) in stroke patients treating intravenous thrombolysis IVT^13^. For AIS patients receiving IVT, elevated PP during is an independent predictor of poor early outcome at hospital discharge and 30-day mortality^16^. Besides, PP is also considered to be closely related to the occurrence^14^ and recurrence^15^ of AIS. However, it is uncertain whether the results of IVT can be extrapolated to patients undergoing EVT with little research on PP for AIS patients after EVT. In this paper, we further explored PP into the analysis of the relationship between post-procedural BP parameters within 24h and primary and second outcomes after thrombectomy.

## METHODS

### Study design and populations

This is a retrospective analyzing consecutive AIS patients with large vessel occlusion who underwent EVT from March 2016 to August 2023 in the Stroke Center at the Department of the Second Affiliated Hospital, Zhejiang University (**Figure 1**). We included patients based on the following criteria: (1) patients who underwent thrombectomy employing second-generation stent-retriever devices or aspirator (Solitaire AB/FR, Covidien/ev3, Irvine, CA; Trevo Proview, Stryker, CA); (2) patients with occlusion of the large artery defined by digital subtraction angiography (DSA), including internal carotid artery (ICA), middle cerebral artery (MCA M1; M2), basilar artery(BA) and vertebral plus basilar artery (VPBA); (3) modified Rankin Scale (mRS) score before the index stroke≤1; (4) EVT could be administered within 6 hours after symptom onset or within 6–24 hours after symptom onset with the presence of large ischemic mismatch/penumbra according to CT perfusion^17^.

**Figure 1.**
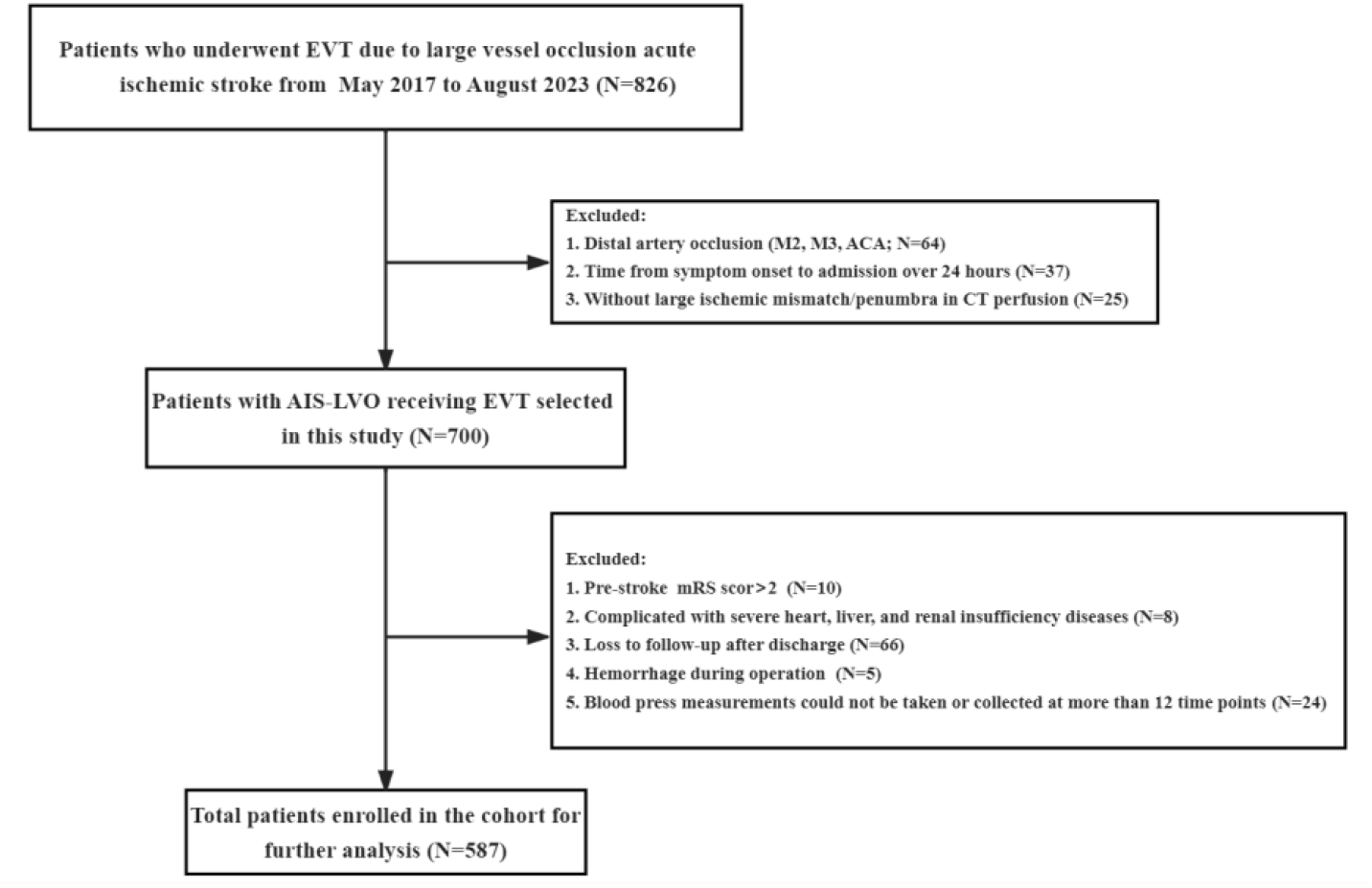
Flow chart of enrolled patients. EVT, endovascular thrombectomy; ICA, internal carotid artery; IVT, intravenous thrombolysis; ACA, anterior communicating artery; M2/M3, second/third segment of the middle cerebral artery; mRS, modified Rankin Scale.

The exclusion criteria were as follows: (1) presence of intracranial hemorrhage identified on CT scan prior to EVT; (2) severe cardiac, hepatic, or renal insufficiency, or advanced diabetes mellitus with blood glucose levels exceeding 22 mmol/L; (3) Over 20% of postoperative blood pressure records were unavailable due to post-operative examination, treatment, or other conditions; (4) lack of follow-up imaging examinations or mRS scores after 3 months post-EVT. This rigorous selection process ensures a homogeneous study population that is representative of clinical practice while minimizing confounding factors that could influence the study outcomes.

### Blood pressure monitoring and measurement

Trained research coordinators documented and collected baseline demographics, clinical characteristics, stroke characters, and imaging data for all participants. Non-invasive BP monitoring was continuously conducted for patients post-EVT. Systolic blood pressure (SBP) and diastolic blood pressure (DBP) measurements were recorded at 15 minutes intervals within the initial 2 hours post-EVT, 30 minutes intervals between 3 and 6 hours, and 1 hour intervals between 7 and 24 hours. Following the guidance of AIS, the selection of antihypertensive medications was at the discretion of the treating clinicians, with calcium channel blockers being the most prescribed.

We measured and calculated the following parameters: (1) mean SBP (average SBP within 24h post-procedure); (2) mean DBP (average DBP within 24h); (3) mean PP (average PP within 24h; mean SBP - mean DBP); (4) maximum SBP (reflecting peak in SBP); (5) maximum DBP (reflecting peak in DBP); (6) minimum SBP (reflecting drop in SBP); (7) minimum DBP (reflecting drop in DBP); (8) SBP-DMM (difference between maximum and minimum in SBP within 24h); (9) DBP-DMM (difference between maximum and minimum in DBP within 24h).

### Follow-up and outcomes

The follow-up protocol was consistent with previously published literatures^18^. The primary outcome measure was functional outcome according to the modified Rankin Scale, which is a 7-point scale ranging from 0 no symptoms to 6 death, assessed at 90 days after EVT^18^. The scores were collected by a stroke neurologist during routine follow-up visits at 90 days (±14) after stroke for the majority of patients through telephone discussion or clinical follow-up with patients or their families. Second outcome was defined as morality at discharge, 3 months and 12 months and sICH. HT referred to the bleeding caused by the reperfusion of blood vessels in the ischemic area after acute cerebral infarction^19^. sICH was defined as any intracranial hemorrhage with an increase in the NIHSS score of≥4 from baseline) within 7days after EVT, according to the ECASS (European--Australasian Acute Stroke Study) II criteria^19^.

### Statistical Analysis

Patients were dichotomized by favorable or unfavorable outcomes groups based on mRS score at 3 months after EVT were 0-3 or 4-6. For missing data sets, we use multiple interpolation to fill in the missing values. Characteristics are summarized as proportions for categorical variables and mean ± SD or median (25-75th percentile) for quantitative variables, as appropriate. Two-sample t-tests or Mann-Whitney U test were used to compare the dichotomous in continuous quantitative variables, while χ2 tests or Fisher’s exact test were for categorical variables. Through ANOVA and univariate logistic regression, potential nine risk BP parameters were filtrated to be associated with primary and second outcomes of AIS patients (P<0.05), then these potential predictors were corrected by confounding factors which were filtrated from clinical characters through univariate logistic regression (P<0.05). After adjusting confounding factors, four variables (mean SBP, mean PP, maximum SBP and SBP-DMM) were filtrated to be independent predictors of short and long-term prognosis. After correcting the above four risk factors at the same time, mean PP was the only independent risk factor and predictor of poor prognosis. Above results were displayed as adjusted odds ratios (OR) combined with a 95% CI. The dose-response relationship between mean PP during hospitalization and adverse outcomes of ischemic stroke was evaluated using restricted cubic splines (RCS). The ROC was used to determine the cut-off value of predictors and their accuracy for the prediction of short-medium prognosis. Subgroup analysis was conducted to further analyze the different prognostic efficacy of coagulation function in certain subgroups. P values of <0.05 were considered statistically significant. All reported P values were two-sided. All statistical analyses were performed using IBM SPSS Statistics for Windows version 26 and R language 4.0.5.

## Results

### Baseline characters

Among 587 patients finally enrolled in the cohort, 326 (55.54%) had an unfavorable 3-months prognosis after EVT who were unable to walk independently. The characteristics of the patients are shown in **Table 1**. Among the cohort, the patients’ mean±SD age was 69.02±13.30 years, with 333 (56.73%) being male and a median admission mRS score of 4. Patients with poor prognosis had a significantly higher mean age, time from puncture to reperfusion (PTR), baseline NHISS and mRS score on admission and with higher proportion of female, hypertension, CHD, diabetes, atrial fibrillation, general anesthesia (unable to cooperate with agitation), difficult thrombectomy (greater than three times). Conversely, higher proportion of successful reperfusion (eTICI = 2B or 3) and MCA occlusion but lower proportion of VERT/BA occlusion was observed in patients with favorable prognosis. Then, above significant changing variables were adjusted for analysis of the association between BP variables and primary and second outcome after EVT.

**Table 1.**
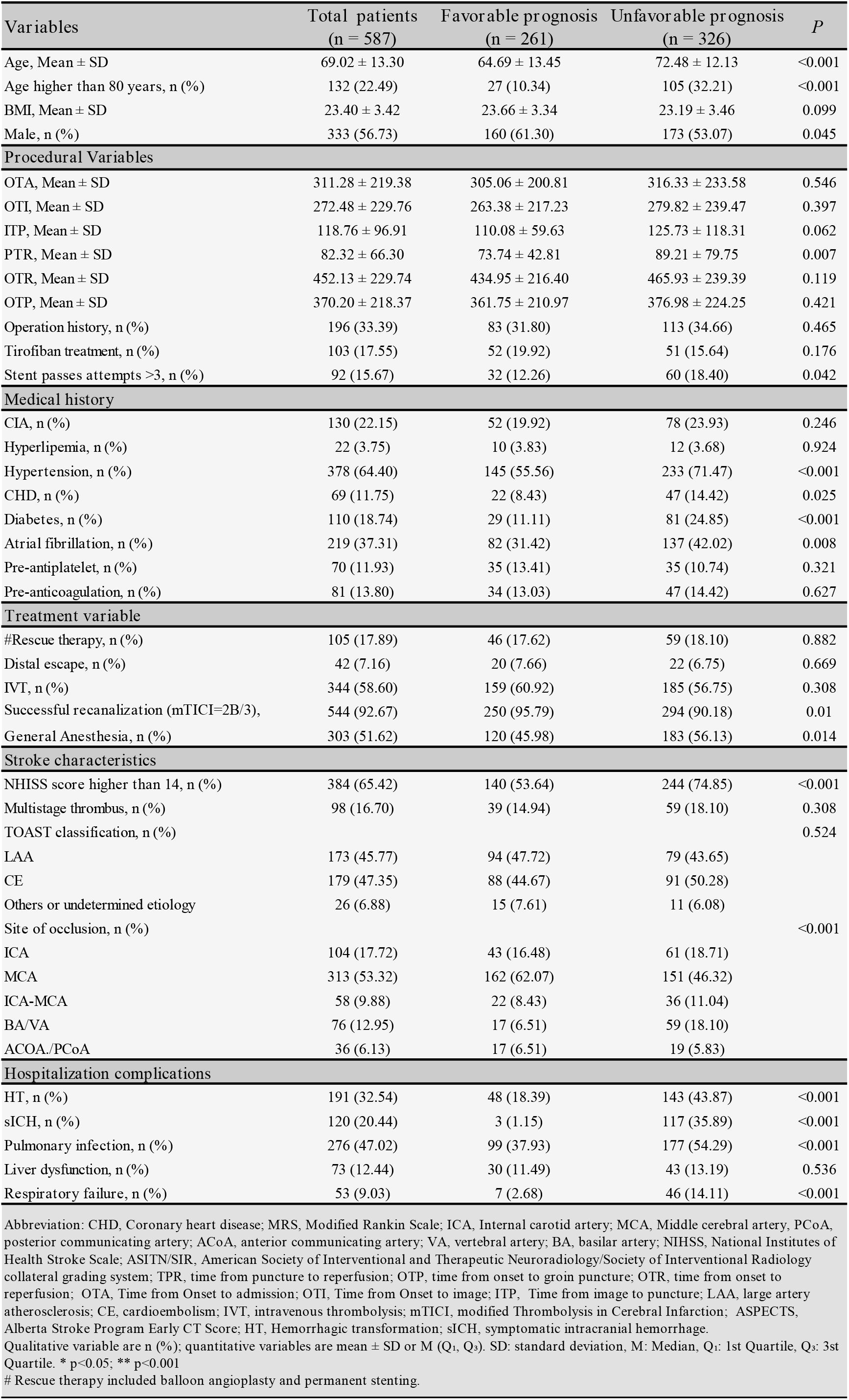
Baseline characteristics of patients with favorable or unfavorable outcomes after 3 months for endovascular treatment of acute large artery occlusive ischemic stroke.

Nine BP parameters were significantly different between two different prognosis. Post-procedural SBP parameters (including baseline SBP, mean SBP, maximum SBP, and SBP-DMM) were significantly higher in patients with poor functional outcomes than in those with good functional outcomes. Inversely, patients with unfavorable prognosis exhibited lower post-procedural DBP parameters (including mean DBP, minimum DBP, and DBP-DMM) than patients with favorable prognosis. As a parameter reflecting dynamic changes in BP, patients with unfavorable prognosis presented higher mean PP.

### Association of Blood pressure variables with primary and second outcomes after EVT

We have adjusted for eleven confounding factors with a significant difference of *P*<0.05 in **Table 2 and Online supplementary Table 1 and 2**. In short-term prognosis, mean SBP (aOR=1.02; 95%CI 1.01 to 1.04), maximum SBP (aOR=1.02; 95%CI 1.01 to 1.03), SBP-DMM (aOR=1.02; 95%CI 1.01 to 1.03), and mean PP (aOR=1.04; 95%CI 1.02 to 1.06) within 24h post-procedural were significantly associated with unfavorable functional outcomes. In long-term prognosis, in addition to the above four variables, DBP-DMM (aOR=1.02; 95%CI 1.01 to 1.04), and minimum DBP (aOR=0.98; 95%CI 0.96 to 0.99) were significantly associated with unfavorable functional outcomes. Mean SBP, mean PP, maximum SBP and SBP-DMM are both independently associated with functional outcomes at 3 and 12 months, in which the correlation of mean PP to prognosis was the strongest with the highest ORs. Then, we further explored the association between above four variables and morality (at discharge, 3 months and 12 months) and sICH in **Online supplementary Table 3**. We found that mean PP was the independent risk factor and predictor of morality at admission (aOR=1.07; 95%CI 1.02 to 1.12), morality at 3 months (aOR=1.02; 95%CI 1.01 to 1.04), morality at 12 months (aOR=1.03; 95%CI 1.01 to 1.04) and sICH (aOR=1.02; 95%CI 1.01 to 1.04) with the highest ORs than other three variables.

**Table 2.** Unadjust and adjusted ORs of the association of post-procedure blood pressure variables with unfavorable functional prognosis at 3 and 12 months after endovascular treatment.

| Blood pressure variables | Unfavorable functional prognosis at 3 months |  |  |  |  | Unfavorable functional prognosis at 12 months |  |  |  |  |
| --- | --- | --- | --- | --- | --- | --- | --- | --- | --- | --- |
|  | Univariate |  | Multivariate |  |  | Univariate |  | Multivariate |  |  |
|  | OR (95% CI) | P | aβ | aOR (95% CI) | aP | OR (95% CI) | P | aβ | aOR (95% CI) | aP |
| Mean SBP | 1.03 (1.02 ~ 1.05) | <0.001 | 0.02 | 1.02 (1.01 ~ 1.04) | 0.005 | 1.03 (1.02 ~ 1.05) | <0.001 | 0.02 | 1.02 (1.01 ~ 1.04) | 0.011 |
| Mean DBP | 0.97 (0.96 ~ 0.99) | 0.005 | -0.02 | 0.98 (0.96 ~ 1.00) | 0.08 | 0.98 (0.96 ~ 0.99) | 0.017 | -0.01 | 0.99 (0.96 ~ 1.01) | 0.182 |
| Mean PP | 1.05 (1.03 ~ 1.07) | <0.001 | 0.04 | 1.04 (1.02 ~ 1.06) | <0.001 | 1.05 (1.03 ~ 1.06) | <0.001 | 0.03 | 1.03 (1.01 ~ 1.05) | <0.001 |
| Maximum SBP | 1.03 (1.02 ~ 1.04) | <0.001 | 0.02 | 1.02 (1.01 ~ 1.03) | <0.001 | 1.03 (1.02 ~ 1.04) | <0.001 | 0.02 | 1.02 (1.01 ~ 1.04) | <0.001 |
| Maximum DBP | 1.00 (0.98 ~ 1.01) | 0.549 |  |  |  | 1.00 (0.99 ~ 1.01) | 0.968 |  |  |  |
| Minimum SBP | 1.00 (0.99 ~ 1.02) | 0.462 |  |  |  | 1.00 (0.99 ~ 1.01) | 0.885 |  |  |  |
| Minimum DBP | 0.96 (0.95 ~ 0.98) | <0.001 | -0.03 | 0.97 (0.95 ~ 1.00) | 0.075 | 0.97 (0.95 ~ 0.99) | 0.002 | -0.02 | 0.98 (0.96 ~ 0.99) | 0.041 |
| SBP-DMM | 1.02 (1.01 ~ 1.03) | <0.001 | 0.02 | 1.02 (1.01 ~ 1.03) | <0.001 | 1.03 (1.02 ~ 1.04) | <0.001 | 0.02 | 1.02 (1.01 ~ 1.04) | <0.001 |
| DBP-DMM | 1.02 (1.01 ~ 1.03) | 0.027 | 0.01 | 1.01 (1.00 ~ 1.03) | 0.105 | 1.02 (1.01 ~ 1.03) | 0.013 | 0.02 | 1.02 (1.01 ~ 1.03) | 0.04 |

Adjusting for confounders included the baseline and procedural variables with a significant difference of $P < 0.05$ (male, age higher than 80 years, successful recanalization (mTICI=2B or 3), general anesthesia, NIHSS higher than 14, hypertension, CHD, diabetes, site of occlusion, atrial fibrillation and number of pass time over 3).
Abbreviation: SBP, systolic blood pressure; DBP, diastolic blood pressure; PP, pulse pressure; DMM, difference between maximum and minimum.
OR: Odds Ratio, CI: Confidence Interval. \* $p < 0.05$ ; \*\* $p < 0.001$ .

**Table 3.** Adjusted ORs of the association of elevated mean PP (>57.39mmHg) post-procedure with pr imary and second outcome events after endovascular tr eatment.

| Outcome events | Univariate |  | Multivariate<sup>&</sup> |  |  |
| --- | --- | --- | --- | --- | --- |
|  | OR (95% CI) | P | aβ | aOR (95% CI) | aP |
| Primary outcome |  |  |  |  |  |
| Unfavorable prognosis at 3 months | 3.08 (2.20 ~ 4.33) | <0.001 | 0.87 | 2.39 (1.58 ~ 3.62) | <0.001 |
| Unfavorable prognosis at 12 months | 2.76 (1.97 ~ 3.86) | <0.001 | 0.73 | 2.08 (1.37 ~ 3.14) | <0.001 |
| Second outcome |  |  |  |  |  |
| Mortality at discharge | 9.49 (2.18 ~ 41.27) | 0.003 | 2.08 | 8.00 (1.68 ~ 38.12) | 0.009 |
| Mortality at 3 months | 1.88 (1.29 ~ 2.75) | 0.001 | 0.42 | 1.52 (0.99 ~ 2.33) | 0.058 |
| Mortality at 12 months | 1.99 (1.40 ~ 2.82) | <0.001 | 0.51 | 1.66 (1.11 ~ 2.50) | 0.014 |
| Symptomatic intracerebral hemorrhage | 1.97 (1.30 ~ 2.98) | 0.001 | 0.47 | 1.60 (1.01 ~ 2.55) | 0.048 |

<sup>&</sup> Adjust variables of outcome after 3 months incorporate PTR, male, age higher than 80 years, successful recanalization (mTICI≥2B/3), general anesthesia, HT, sICH, pulmonary infection, respiratory failure, respiratory failure, NHISS higher than 14, hypertension, CHD, diabetes, site of occlusion, atrial fibrillation and number of sent pass over three times. Abbreviation: SBP, systolic blood pressure; DBP, diastolic blood pressure; PP, pulse pressure; PTR, time from puncture to reperfusion. OR: Odds Ratio, CI: Confidence Interval. \* p<0.05; \*\* p<0.001.

### PP may be a better predictor of outcomes after EVT than SBP

We used the receiver operating characteristic curve (ROC) to further validate the diagnostic efficacy of the above four variables in **online Figure 1**. Among them, mean PP within 24h post-procedure had the optimal diagnostic efficacy on poor outcome after EVT with the cut-off value of 57.39 mmHg, presenting a sensitivity of 62.7% and a specificity of 65.1% with the AUC of 0.661 (95% CI 0.617 to 0.705). When we corrected the above four risk factors at the same time, mean PP was the only independent risk factor and predictor of poor prognosis at 3 months (aOR=1.04; 95%CI 1.01 to 1.07) and 12 months(aOR=1.03; 95%CI 1.01 to 1.06), morality at discharge (aOR=1.09; 95%CI 1.02 to 1.17), and morality at 3 months (aOR=1.04; 95%CI 1.01 to 1.06) in **supplemental online Table 4 and 5**.

Restricted cubic spline (RCS) with 3 knots was used to explore the relationship between postoperative PP and outcome variables after adjusting confounding factors in **Figure 2**. Mean PP demonstrates a significant dose-response relationship with the occurrence of functional outcomes, sICH, and mortality after thrombectomy, with an increased risk of adverse outcomes as PP rises. Besides, mean PP exhibits a linear relationship with all other outcome events, except for mortality at 12 months post-EVT.

**Figure 2A.**
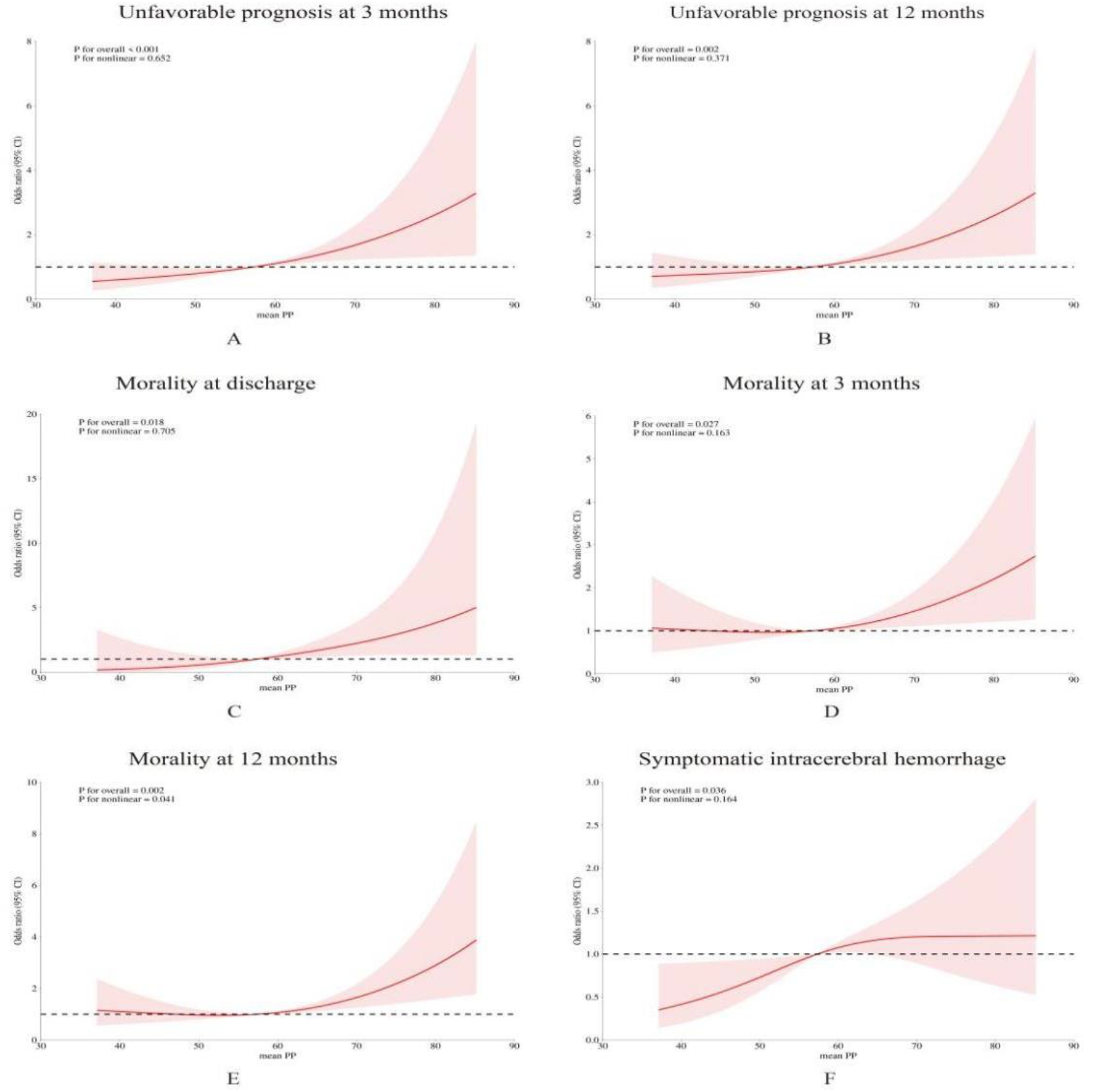
The dose-response relationship between postoperative mean PP and unfavorable prognosis at 3 months through restricted cubic splines (RCS) with 3 knots. Figure 2B. The dose-response relationship between postoperative mean PP and unfavorable prognosis at 12 months. Figure 2C. The dose-response relationship between postoperative mean PP and morality at discharge. Figure 2D. The dose-response relationship between postoperative mean PP and morality at 3 months. Figure 2E. The dose-response relationship between postoperative mean PP and morality at 12 months. Figure 2F. The dose-response relationship between postoperative mean PP and symptomatic intracerebral hemorrhage.

### Association of elevated mean PP with primary and second outcomes after EVT

Then, based on the cut-off value determined by RCS, the continuous mean PP variable was categorized into groups above 57.39 mmHg and below 57.39 mmHg. Compared with patients with lower mean PP (<57.39mmHg), patients with higher mean PP within in 24 hours following EVT were more likely to complicated sICH (aOR=1.06; 95%CI 1.01 to 2.55) and have worse functional outcomes at 3 months and 12 months (aOR=2.39; 95%CI 1.58 to 3.62; aOR=2.08; 95%CI 1.37 to 3.14) or higher risk of morality at discharge (aOR=8.00; 95%CI 1.68 to 38.12) and at 12 months after EVT (aOR=1.66; 95%CI 11 to 2.50) in **Table 3 and supplemental online Table 4**.

### Subgroup analysis of post-procedure PP with functional outcomes

Subgroup variables were selected from positive clinical risk factors after multivariate correction. In most subgroups, higher PP group (>57.39 mmHg) was significantly associated with poor functional outcomes at 3 and 12 months after EVT in **Figure 3**. Regardless of patients’ gender, type of anesthesia, presence or absence of comorbidities such as hypertension and diabetes, and whether the number of thrombectomies exceeds three times, controlling PP below 57.39mmHg is beneficial for improving prognosis. However, there is no significant interaction effects between PP custom and various stratification factors on outcomes at 3 months post-thrombectomy (all P>0.05). In term of outcome at 12 months, we unexpectedly found that in the subgroup characterized by more than three thrombectomy procedures, age less than 80 years, and anterior circulation infarction, the benefit of maintaining PP below 57.39 mmHg is significantly greater than that of its corresponding subgroup.

**Figure 3A.**
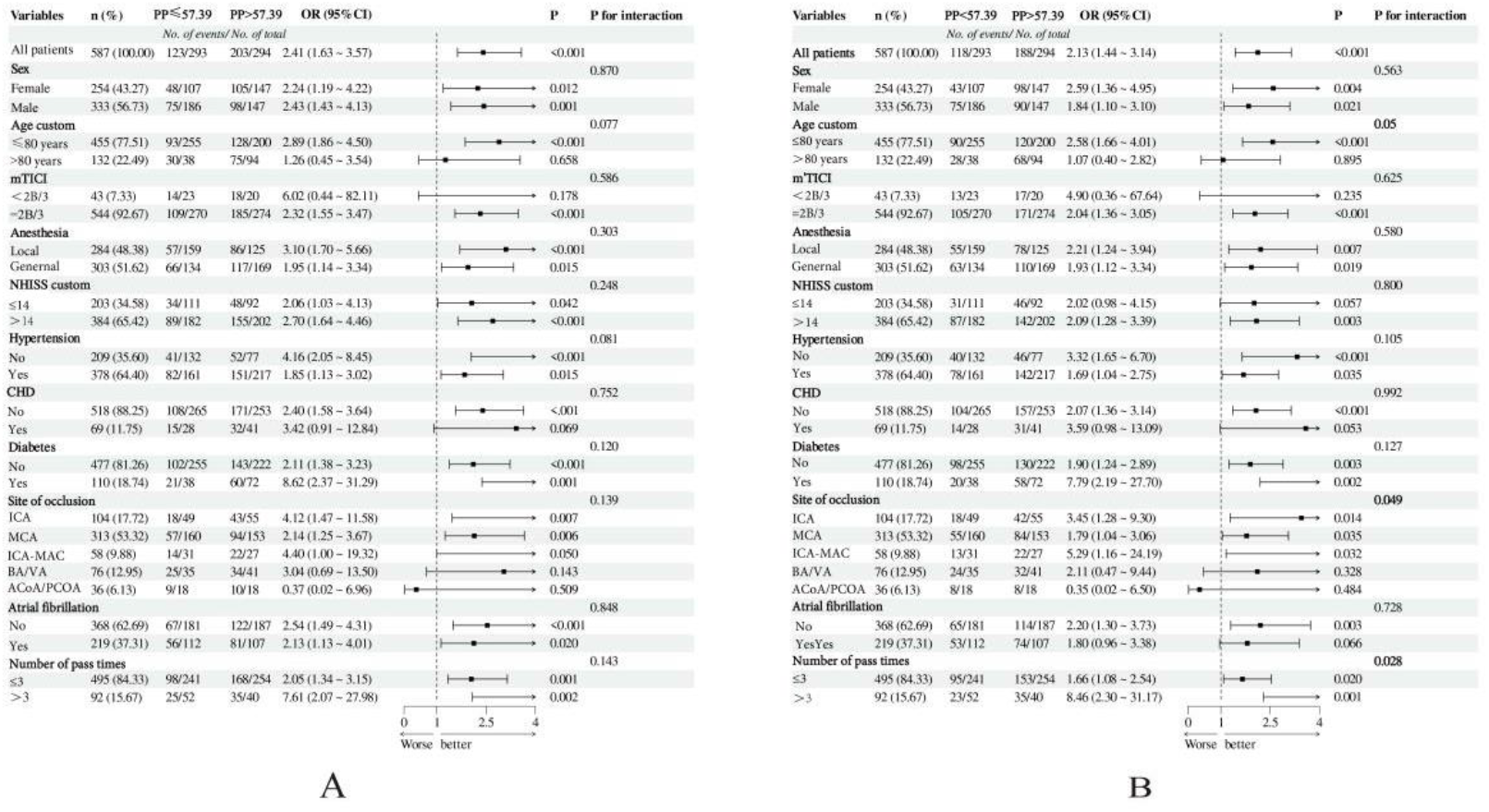
Subgroup analysis of patients with 90-day modified Rankin Scale (mRS) score of 0–3. Figure 3B. Subgroup analysis of patients with 1-year modified Rankin Scale (mRS) score of 0–3.

## Discussion

According to our study, we found that mean SBP, mean PP, maximum SBP and SBP-DMM were both positively significantly associated with unfavorable prognosis at 3 and 12 months. Among them, mean PP present RCS analysis indicated that mean PP exhibits a linear relationship with all other outcome events, except for mortality at 12 months post-EVT. The predictive power and strength of association of mean PP with prognosis are superior to those of single SBP or DBP alone with the strongest ORs and highest diagnostic performance (AUC=0.661, 95% CI 0.617 to 0.705). Mean PP within 24 hours was closely associated with primary and second outcomes, which was not only an independent risk factor and predictor of prognosis, but also an independent risk factor for morality and sICH after EVT. We also found that in the subgroup characterized by more than three thrombectomy procedures, age less than 80 years, and anterior circulation infarction, the benefit of maintaining pulse pressure below 57.39 mmHg is significantly greater than that of its corresponding subgroup.

When ischemic occur, excessive systemic BP may lead to a breach in cerebral perfusion pressure, resulting in hemorrhagic transformation, while too low systemic pressure may cause inadequate perfusion pressure in a decline in collateral compensation capacity, further exacerbating the infarction^20^. Therefore, balancing the risks of intracranial hyperperfusion and hypoperfusion by optimal post-procedure peripheral BP and perfusion pressure is an important part of the treatment during hospitalization for patients with AIS^21^. American Heart Association/American Stroke Association (AHA/ASA) guidelines recommend a fixed blood pressure target of ≤ 180/105 mmHg during and for 24 h after the procedure (IIa recommendation)^17^. However, the above recommendations are mainly based on the relevant evidence of intravenous thrombolysis (IVT). At present, there is limited data to guide BP management for AIS-LVO patients receiving EVT, and no unequivocal consensus exists regarding the optimal BP target before, during, and after the EVT procedure. Despite recently completed several clinical trials on BP managements post-EVT, the conclusions remain inconsistent.

Safety and efficacy of intensive blood pressure lowering after successful endovascular therapy in acute ischemic stroke (BP-TARGET) was the first RCT to investigate BP management after mechanical thrombectomy, which showed that intensive BP lowering group (100 to 129 mmHg) did not significantly reduce the risk of spontaneous hemorrhagic transformation than control group (130 to 185 mmHg), and there was no statistically significant difference in clinical prognosis between the two groups (aOR=0.96, 95%CI 0.60 to 1.51, *P*=0.84)^22^. The OPTIMAL-BP study incorporated a total of 306 participants, which obtained a similar result to the ENCHANTED2/MT[13]. The positive prognosis (mRS=0 to 2) at 3 months after surgery in the intensive antihypertensive group (SBP<140 mmHg) was worse than that in the control group (SBP=140 to 180 mmHg) (39.4% vs 54.4%; aOR=0.56, 95% CI 0.33 to 0.96), which was terminated early due to safety concerns. The ENCHANTED2/MT study is designed to study whether intensive antihypertensive therapy (SBP< 120 mmHg) could improve functional outcomes in patients over higher BP management strategies (SBP=140 to 180 mmHg) in patients with AIS-LVO who have successfully recanalized after EVT^23^. The large RCT intended to include 2257 participants and was terminated early in the interim analysis due to reaching the safety endpoint ahead, eventually ended with 821 participants, demonstrated for the first time that a hypertensive strategy with a SBP< 120 mmHg was harmful for patients with AIS-LVO after successfully recanalization (aOR=1.37, 95% CI 1.07 to 1.76), verifying the lower limit of safety management^23^. The optimal BP target to avoid both postoperative hemorrhagic transformation and cerebral perfusion injury remains unknown.

Pulse pressure (PP) is an important hemodynamic parameter, representing the difference between the maximum and minimum value of circulating pressure in a cardiac cycle^25^. Determined by the interplay of cardiac stroke volume, systemic arterial system, and peripheral microcirculatory pressure, high PP may reflect impairment of the body’s blood pressure autoregulatory function^26^. In previous studies, it has been reported that higher PP at admission was closely related to the occurrence and recurrence of AIS [16]. A cohort study involving 9,901 patients with hypertension demonstrated that compared to patients in the lowest quartile (<50 mmHg), those in the highest quartile (>74 mmHg) had a significantly increased risk of occurrence of AIS (HR: 1.555, 95%CI: 1.127-2.146) [16]. Another study incorporating 1009 young ischemic stroke patients indicated that PP also was a risk factor for recurrence of AIS, which demonstrated that patients with higher admission PP had a higher risk of stroke recurrence (HR: 1.11, 95%CI: 1.01 to 1.21)^15^. High PP was also an independent risk factor for the development of AIS, with the highest quartile (>74mmHg) at a significantly increased risk than the lowest quartile (<50mmHg) (HR: 1.56, 95%CI: 1.13 to 2.15)^14^. Another study involving 674 patients with AIS treated with IVT found that higher PP and fluctuations were significantly associated with poor neurological function within 24 hours and adverse functional outcomes at 90 days, and were independently associated with the risk of death within 90 days (OR: 1.60, 95% CI: 1.23-2.07), suggesting that monitoring and management of PP are crucial in the treatment and prognosis evaluation of AIS^27^.

It is unknown whether the PP after thrombectomy will have an impact on prognosis. We demonstrated that mean PP post-procedure is not only an independent predictor of outcomes at 3 and 12 months post-thrombectomy but is also closely associated with symptomatic intracranial hemorrhage and postoperative mortality. The predictive power and strength of association of PP with prognosis are superior to those of SBP or DBP alone, with the highest diagnostic performance and ORs. The mechanism by which reducing PP improves the prognosis of stroke patients and reduces the incidence of sICH is not yet clear. The possible mechanism is that the ischemic penumbra is highly sensitive to blood pressure changes and excessive fluctuations after the onset of ischemic stroke, aggravating local ischemia, and in turn, leading to poor prognosis^28^. In addition, PP may also be associated with cerebrovascular lesions and brain white matter damage in ischemic condition, which can further lead to cognitive and other disorders. Elderly people with increased PP have a greater risk of neuronal injury, and reducing PP may improve mRS scores through slowing down progressive cerebrovascular injury^29^. In the subgroup characterized by more than three thrombectomy procedures, age less than 80 years, and anterior circulation infarction, the benefit of maintaining PP below 57.39 mmHg is significantly greater than that of its corresponding subgroup, which may help us further identify the optimal population for controlling PP.

Our study also found that mean SBP, maximum SBP and SBP-DMM were associated with prognosis of AIS after EVT, which was similar to previous studies^30,31^. An observational multicenter study suggested that patients with post-procedural SBP targets of <140mmHg and <160 mmHg showed a significantly higher proportion of good functional outcomes, compared with the guideline-recommended SBP of <180 mmHg^31^. Furthermore, a 24-hour mean SBP of <140 mmHg post-EVT was associated with functional independence, while a higher SBP was independently correlated with sICH, mortality and the requirement for hemicraniectomy^30^. A meta-analysis including seven RCTs (MR CLEAN, ESCAPE, EXTEND-IA, SWIFT PRIME, REVASCAT, PISTE, THRACE) indicated that SBP was associated with worse clinical outcomes in patients with baseline SBP≥140 mmHg (aOR 0.86, 95% CI 0.81-0.91). There was no significant association for patients with baseline SBP < 140 mmHg^24^. High maximum SBP levels following MT are independently associated with increased likelihood of 3-month mortality and functional dependence in LVO patients. Moderate BP control is also related to lower odds of 3-month mortality in comparison to permissive hypertension^33^. In fact, most scholars still insist that strengthening BP reduction is correct, but the timing of BP reduction and the speed and degree of depressurization, which still needs more further studies.

## Limitations

Our study has several limitations. First, due to the retrospective observational design, results could have been confounded by variables not adjusted for in the analyses, so residual confounding might be present. Second, as we did not have data on individual SBP targets or information on administration of either a vasopressor or an antihypertensive agent after EVT, we do not know how well SBP was managed. Thirdly, since the cohort included patients who did not achieve posterior circulation and did not achieve complete vascularization, further group analysis of patients may help to more accurately assess the effect of systolic blood pressure on prognosis. The 24-hour post-procedural SBP fluctuation may be a result of BP management or the natural course of ischemic stroke. Finally, another limitation to note is that BP monitoring is intermittent, so fluctuations between cuff pressure checks are not recorded. Therefore, further research is warranted for validation.

## Conclusions

Mean PP within 24 hours after EVT present a significant dose-response relationship with the occurrence of functional outcomes, sICH, and mortality after thrombectomy, with an increased risk of adverse outcomes as PP rises. Besides, mean PP exhibits a linear relationship with all other outcome events, except for mortality at 12 months post-EVT. Controlling mean PP below 57.39 mmHg is beneficial to improve prognosis, especially in patients who younger than 80 years, more than three thrombectomy procedures, age less than 80 years, and anterior circulation infarction. Therefore, for patients with AIS-LVO, BP management should not only focus on reducing SBP levels but also on stabilizing PP fluctuations. Achieving consistent long-term control of SBP and PP may be beneficial for improving the prognosis of ischemic stroke patients, which is needed to be verified new large-scale randomized controlled trials (RCTs).

## Data Availability

Funder requirements do not apply, but at the time of publication, the data will be available in an article or appropriate repository.

## Declarations

### Ethical Approval and Consent to participate

This large retrospective study was approved by human Research Ethics Committee, the Second Affiliated Hospital of Zhejiang University School of Medicine. Without causing any potential harm to the patient, the patient’s informed consent is exempt.

### Consent for publication

Not applicable.

### Availability of data and materials

Data and material are not publicly available, but can be requested.

### Competing interests

The authors declare that the research was conducted in the absence of any commercial or financial relationships that could be construed as a potential conflict of interest.

### Funding

This work was financially supported by the National Natural Science Foundation of China (82202407), Natural Science Foundation of Zhejiang Province (LQ21H090008) and Zhejiang medical and health science and technology project (2021434317).

### Authors’ contributions

Conception and design: J.L. and S.J.

Acquisition of data: C.Q. and J.Y. and X.L.

Analysis and interpretation of data: L.X.

Drafting of the article: S. J. and P.Z. and Y.Y.

Critically revising the article: F.B. and Z.T.

Reviewed submitted version of manuscript: all authors.

Approved the final version of the manuscript on behalf of all authors: J.L. and J.X.

Administrative/technical/material support: X.C. and F.B.

Study supervision: C.Q

## Acknowledgments

This work was financially supported by the National Natural Science Foundation of China (82202407), Natural Science Foundation of Zhejiang Province (LQ21H090008) and Zhejiang medical and health science and technology project (2021434317). We also thank Lingzhi Yu for her comments on the manuscript.

## Notes

### Competing Interest Statement

The authors have declared no competing interest.

### Author Declarations

This large retrospective study was approved by human Research Ethics Committee, the Second Affiliated Hospital of Zhejiang University School of Medicine.

